# Labeling self-tracked menstrual health records with hidden semi-Markov models

**DOI:** 10.1101/2021.01.11.21249605

**Authors:** Laura Symul, Susan Holmes

## Abstract

Globally, millions of women track their menstrual cycle and fertility via smartphone-based health apps, generating multivariate time series with frequent missing data. To leverage data from self-tracking tools in epidemiological studies on fertility or the menstrual cycle’s effects on diseases and symptoms, it is critical to have methods for identifying reproductive events, *e*.*g*. ovulation, pregnancy losses or births. We present two coupled hidden semi-Markov models that adapt to changes in tracking behavior, explicitly capture variable– and state– dependent missingness, allow for variables of different type, and quantify uncertainty. The accuracy on synthetic data reaches 98% with no missing data, 90% with realistic missingness, and 94% accuracy on our partially labeled real-world time series. Our method also accurately predicts cycle length by learning user characteristics. It is publicly available (`HiddenSemiMarkov R` package) and transferable to any health time series, including self-reported symptoms and occasional tests.

## Introduction

Health tracking apps have become increasingly popular and self-tracked health records collected via apps or connected devices are progressively adopted by the scientific community for personalized health or epidemiological research (Topol, 2019). Menstrual cycle and fertility tracking apps are among the most used health apps (Fox & Duggan, 2013). These apps are now used by millions of women worldwide, generating very large datasets of self-reports related to the menstrual cycle and reproductive events. Users of these apps typically report their period bleeding along with physical or psychological symptoms and/or fertility-related body-signs.

These large datasets have already been used to characterize the duration of the menstrual cycle and the follicular (before ovulation) and luteal (after ovulation) phases (Bull et al., 2019; Faust et al., 2019; Symul et al., 2019), to evaluate the association between sexually transmittable infections (STI) and pre-menstrual symptoms (Alvergne et al., 2018) and to evaluate the association between cycle length irregularities and reported symptoms (Li et al., 2019). In addition to these findings, this data indubitably holds additional information on fertility, pregnancy losses and menstrual health in general. This information can be used to tackle scientific challenges and address unanswered questions about the human reproductive biology. For example, this data can be used to evaluate whether seasonal and geographical variations of fertility (Symul et al., 2020) is due to changes in ovulation or loss rates or to study the predictability of mental health variations throughout the menstrual cycle (Eisenlohr-Moul et al., 2019; Pierson et al., 2019) at large scale, in individuals not diagnosed with pre-menstrual dysphoric disorder (PMDD) (*Diagnostic and Statistical Manual of Mental Disorders, 5th Edition*, 2013). Beyond the potential of these large retrospective datasets, apps and/or connected devices provide a scalable way to prospectively collect longitudinal data of menstrual-health related body signs and symptoms for large population size, long period of time and without the requirements of in-person visits to a clinic. The prospective digital collection of data related to fertility and menstrual health provides an opportunity to evaluate the association between the menstrual cycle or reproductive status and other health variable at large scale.

A first challenge in using such self-reported data is the contextualization of each observation within the reproductive timeline of an individual. The interpretation of a reported symptom varies greatly if the individual is pregnant or going through a long anovulatory phase. This contextualization requires the labeling of users’ time series with biologically-relevant states such as “pregnant” or “ovulating”.

Labeling self-tracked datasets can be a challenging process given the multivariate nature of the datasets, the abundance of missing data and the lack of available ground-truth. To our knowledge, there are no available labeled datasets for menstrual self-tracked data. Thus, supervised labeling methods such as Long-Short-Term-Memory (LSTM) models (Hochreiter, 1997; Liu et al., 2019), or Transformers (Vaswani et al., 2017), cannot be used. Fortunately, this lack of available labeled samples is balanced by a good knowledge of the underlying reproductive biology. This knowledge can be translated into statistical priors and inform the design of unsupervised or generative models.

For example, it has been well documented that the cervical mucus properties and quantities are under the control of cycling reproductive hormones (Bigelow et al., 2004; Billings et al., 1972; Moghissi et al., 1972) and that these changes can be observed and reported by app users (Faust et al., 2019; Symul et al., 2019). Body temperature at wake-up has been shown to increase after ovulation and in early pregnancy (Buxton & Atkinson, 1948; Moghissi et al., 1972). Concentration in luteinizing hormone (LH) surges before ovulation (Moghissi et al., 1972; Pauerstein et al., 1978) and this surge can be detected by cheap at-home urine kits (Su et al., 2017). Bleeding patterns, the most obvious body-sign to report into a menstrual-cycle tracking app, is highly correlated with menses (periods), pregnancy losses or births, but light bleeding may also be indicative of ovulation or be reported in early pregnancy (Fraser et al., 2011; Harville et al., 2003).

Hidden state models are appropriate for labeling self-tracked time series because the underlying biological states can be matched to the model’s hidden states. In the medical literature, the menstrual cycle is frequently split into successive phases (menses, early follicular phase, peri-ovulatory phase, early and late luteal phases) and pregnancies are frequently divided into trimesters. Given that these phases have been well characterized, they can be naturally translated into a discrete state model. Each latent state matches one of the menstrual or pregnancy phases. In previous work, hidden Markov models (HMM), the most common discrete hidden state model for time series, have been used to label menstrual-cycle time series (Symul et al., 2019). However, the Markovian property imposes a geometrical distribution for the duration of each state which does not match with the description of the menstrual or pregnancy phases. Hidden Markov models only performed well in labeling individual cycles whose start and ends were already identified and where users had reported enough data to constrain the duration of each phase. Others have proposed cyclic HMM (CyHMM) to recover cycle characteristics from menstrual cycle app data (Pierson et al., 2017). While this framework is successful in identifying cycles, it did not include prior biological knowledge beyond the average cycle length. Consequently, the hidden states can not directly be matched to and interpreted as biological states. Additionally, because that framework assumed cycles with relatively small variations in length, it was suitable for identifying menstrual cycles but not pregnancies or post-partum states, preventing the labeling of such events.

Hidden semi-Markov models (HSMM) allow for non-geometric distribution of state duration, often called “state sojourn” in the semi-Markov context. Discrete hidden state models can accurately and realistically describe reproductive events and thus be used to decode fertility app time series. While hidden semi-Markov models have been used in a large variety of applications, ranging from biological sequence analyses (Yu, 2010) to modeling financial market variations (Bulla & Bulla, 2006; D’Amico et al., 2009), there was, to our knowledge no previous implementation that fulfilled the requirements of our task.

Our contributions are (a) the adaptation of hidden semi-Markov models to decode censored (mobile) (health) multivariate time series, (b) the implementation of these changes in a publicly available R package (HiddenSemiMarkov), (c) the definition of a HSMM describing the reproductive biology and (d) a generative method of coupled HSMMs which accounts for longterm changes in tracking behavior.

We evaluate the performances of our method on two datasets: a real-world dataset and a synthetic dataset with varying amounts of missing data. In addition, we evaluate the ability of our model to learn individuals’ cycle characteristics by quantifying the error on cycle length prediction.

Our data is a de-identified dataset provided by the menstrual cycle and fertility tracking app Kindara (see Materials and Methods). This dataset was composed of the self-tracked logs of 64 long-term users of the app. The features reported by users were (1) their bleeding flow (none, spotting, light, medium, heavy), (2) the consistency of their cervical mucus (none, creamy, egg-white, watery, sticky) and the quantity (little, medium, lots) when it was not missing, (3) their body temperature, in Fahrenheit, and whether they marked their temperature measurement as “questionable”, which is recommended by the app if the value is oddly low or high or if the user did not sleep enough hours, (4) the results of LH tests (positive or negative) and (5) the results of pregnancy tests (Fig 1a). Each of these features can be reported daily by users. However, users do not report all of these features every day and there is a large variability in tracking frequency between users. Missing data are very frequent. The average tracking frequency is just above 50%, which means that on average users open and log a feature in the app approximately every other day, but it may be as low as 16% or as high as 100% for some users (see Table 1). Fig 1b provides two examples of time series logged by app users.

**Table 1:** Tracking behavior of the users in our dataset. The first row provides descriptive statistics of the overall tracking frequency, which is defined as the ratio between the number of days in which the user tracked any feature in the app and the duration (in days) between their first and last log. The next four rows provide feature-specific tracking frequencies, i.e. the ratio between the number of days the user tracked this specific feature and the duration of app use. The last two rows provides descriptive statistics about the duration of the longest continuous stretch of time users did not report any feature (missing) or reported as least one feature every day (tracking).

| metric | mean | median | min | 5th percentile | 95th percentile | max |
| --- | --- | --- | --- | --- | --- | --- |
| Overall tracking frequency | 0.55 | 0.47 | 0.16 | 0.20 | 0.99 | 1.00 |
| Temp tracking frequency | 0.30 | 0.14 | 0.00 | 0.00 | 0.92 | 1.00 |
| Mucus tracking frequency | 0.18 | 0.06 | 0.00 | 0.00 | 0.73 | 0.99 |
| LH test tracking frequency | 0.01 | 0.00 | 0.00 | 0.00 | 0.06 | 0.26 |
| Pregnancy test tracking frequency | 0.00 | 0.00 | 0.00 | 0.00 | 0.02 | 0.04 |
| Longest consecutive missing (days) | 183.45 | 55.00 | 1.00 | 6.60 | 711.80 | 1386.00 |
| Longest consecutive tracking (days) | 333.94 | 141.50 | 7.00 | 10.65 | 1473.50 | 2604.00 |

**Figure 1:**
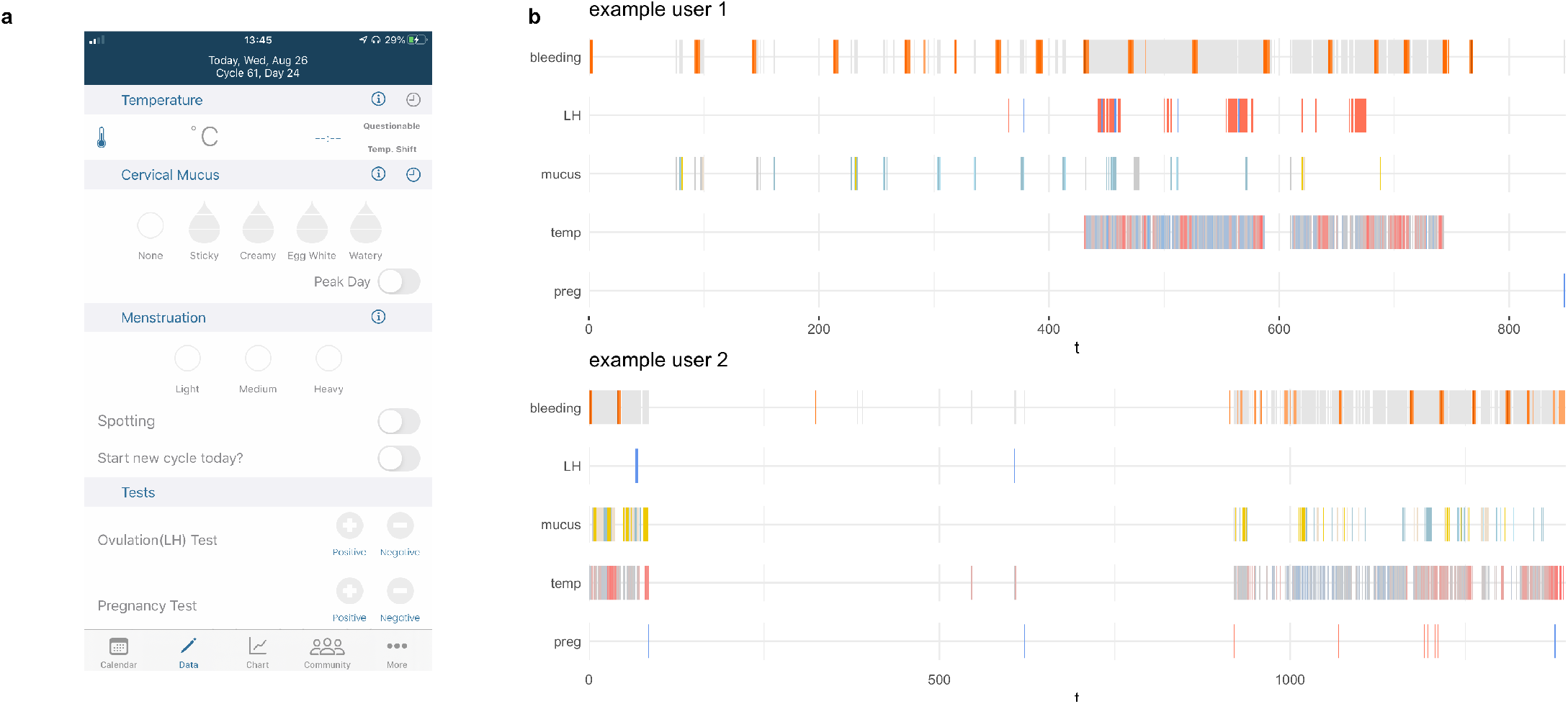
Dataset. (a) Snapshot of the tracking screen of the Kindara app. (b) Examples of time series tracked by two users of the app. For all features, the absence of vertical line indicates missing data for that variable. In the bleeding line (top), gray lines indicate ‘no bleeding’, orange/red lines mean that bleeding was reported. Darker reds indicate heavier bleeding flow. In the LH and pregnancy test lines (2nd and last lines), red lines indicate negative tests, blue lines positive ones. Temperature is depicted by a gradient ranging from blue for temperatures below the user’s median value to red for temperatures above the median value. No mucus (3rd line) is depicted by gray lines, while fertile mucus is indicated in blue, sticky mucus in yellow and creamy one in beige.

Given the generative nature of our model, the synthetic dataset is simulated from our coupled HSMMs. Synthetic time series are generated with various amounts of missing data so that the effect of tracking frequency on accuracy can be evaluated (Methods).

## Results

### Generative models of the female reproductive cycles and of users’ tracking behavior

The self-reported time series result from two distinct generative processes: the first one generates the values of the observed variables and the second one generates the censoring (the pattern of missing data). Here, we hypothesize that the first process only depends on the biological state, while the probability of missing data depends on both the biological state of an individual and on the tracking goals of that individual. For example, the probability of a positive pregnancy test only depends on whether the individual is pregnant (assuming no false positives), however, the probability of *reporting* a positive pregnancy test is much higher in the first month of pregnancy than in the last month of pregnancy. Similarly, a user who wishes to achieve or avoid a pregnancy is likely to report their cervical mucus and/or temperature more frequently than a user whose purpose is simply to keep track of their period. While, in principle, the tracking behavior does not affect an individual’s biology, our ability to detect specific reproductive events depends on the tracking behavior. In general, the more a user tracks, the more information we have about their biological state, and the better specific events can be identified from their data. Inversely, if, for example, a user only tracks their bleeding, it may be impossible to differentiate early pregnancy losses from long cycles.

Consequently, we defined two coupled hidden semi-Markov models: the first one models the female reproductive cycles and the second one models the long-term tracking behavior of users, reflecting their inferred tracking purpose (Fig 2). Together they represent a generative model for the individual’s time series. The specification of these two models is presented in figure 2, and detailed and fully justified in the Methods and the Supplementary Material.

**Figure 2:**
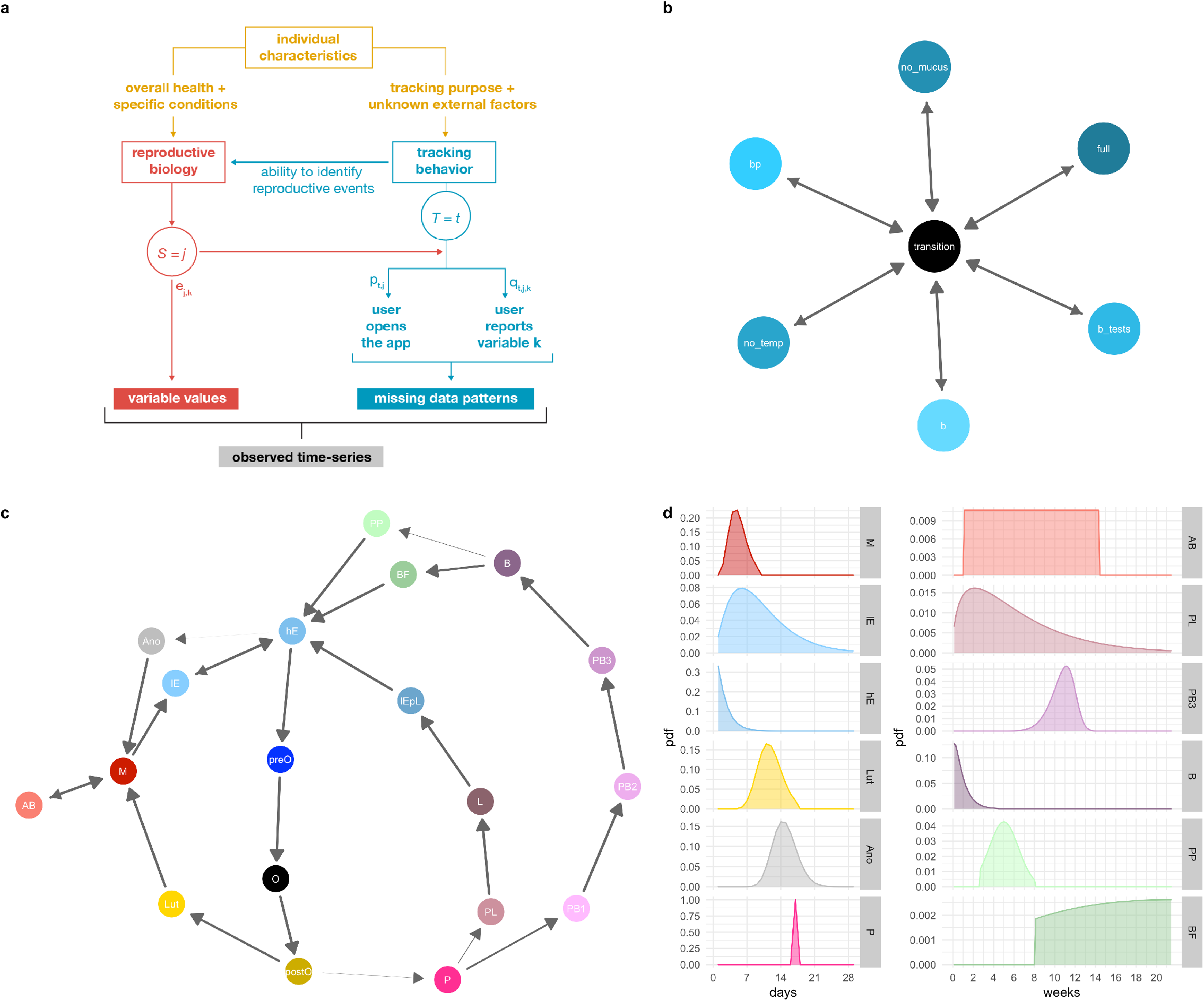
Modeling the data generation process. (a) Graph of the generative model assumed to lead to the observed sequences. (b) Specified model graph of the HSMM used to label users time series with a specific tracking behavior. ‘b’ indicates tracking of bleeding only, ‘bp’ means tracking of bleeding and pregnancy tests, ‘b test’ tracking of bleeding and both ovulation and pregnancy tests, ‘no mucus’ indicates that the user is not tracking their mucus, in ‘no temp’ they are not tracking their temperature and in ‘full’, they are tracking all five possible variables. This does not require them to track these features frequently, but at least occasionally within a few months. (c) Graph of the specified HSMM for modelling reproductive events. Arrows indicate possible transitions, their width is proportional to the transition probability. This graph should be read starting from the red circle (‘M’ for menses). From ‘M’, a first loop matches the 6 states defining ovulatory cycles (‘lE’, ‘hE’, ‘pre-O’ are follicular phase states, ‘O’ stands for ovulation, and ‘post-O’ and ‘Lut’ are luteal phase states). After ovulation, a pregnancy may start (‘P) and end-up in a loss (‘PL’-’L’-’lEpL’ loop) or in a birth (‘PB1’-’PB2’-’PB3’-’B’-’PP’ (post-partum without breast-feeding) or ‘BF’ (breast-feeding) loops). Finally, two anovulatory states are defined: ‘AB’ for anovulatory with bleeding and ‘Ano’ for anovulatory without bleeding. See Methods and Supplementary Material for state definition and descriptions. (d) Prior and initial sojourn distributions for states which do not have a fixed duration.

### Labeling performance on synthetic data

Our results on a synthetic dataset (Fig 3) show that our method is able to recover the ground truth with an accuracy of 98% when variables are always reported (persistent tracking, no missing data). This provides an upper-bound on our ability to decode real-world time series.

**Figure 3:**
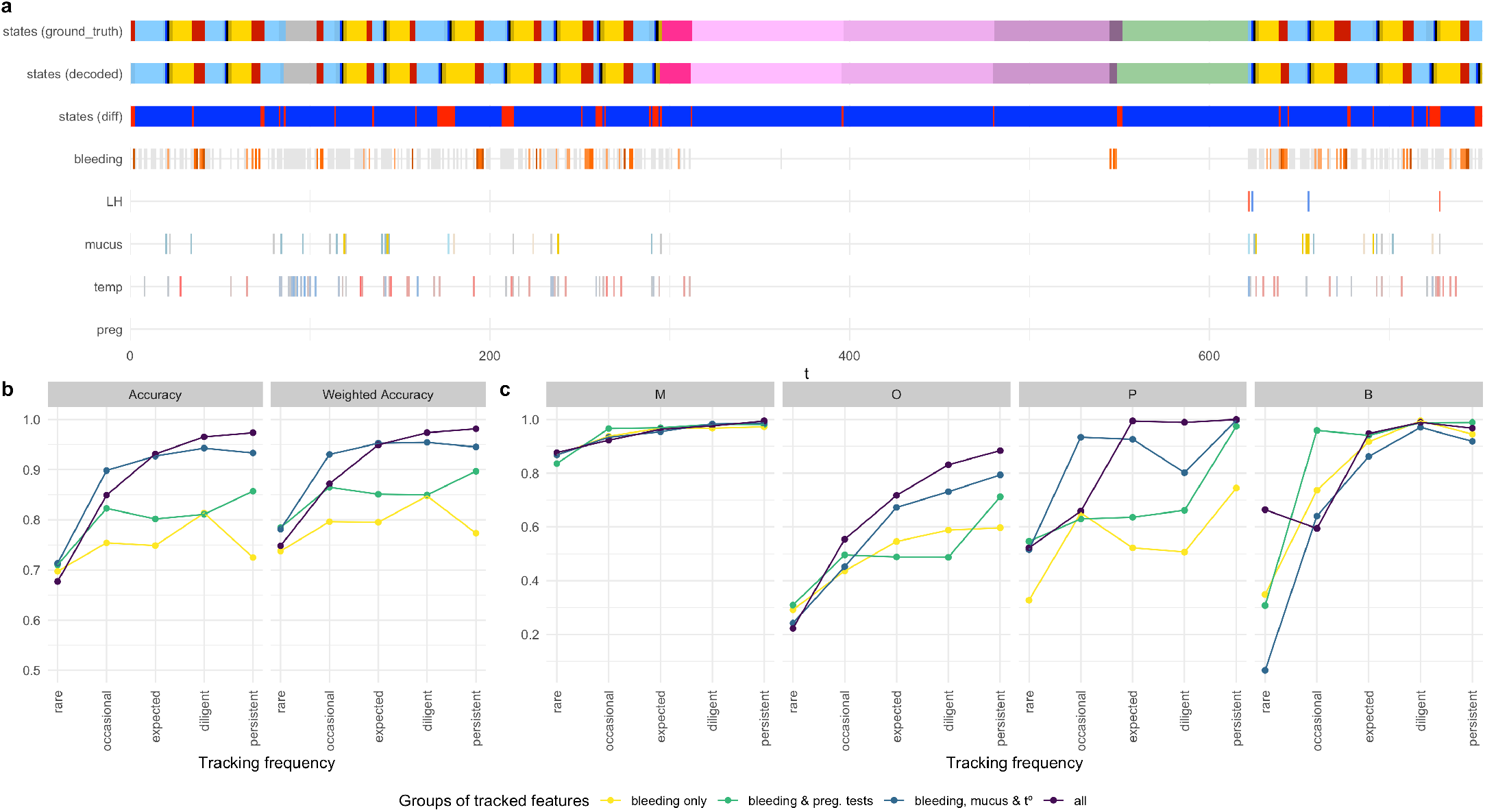
Performance on synthetic dataset. (a) Example of a simulated time series with the ground-truth (i.e. simulated sequence of states, first row) and the sequence of hidden states predicted by our method (second row). The third row shows the difference between the two rows; blue indicates agreement, red indicates labeling errors. (b) Accuracy (left) and weighted accuracy (right) for different tracking frequency (x-axis) and for different sets of tracked variables (colors). (c) State-specific weighted accuracy for the periods (menses - M), ovulation (O), implantation (pregnancies - P) and births (B).

As expected, we observe a higher accuracy when variables are reported more frequently (less missing data, see tracking categories on the x-axis of Fig. 3b-c) and when more variables are reported, *e*.*g*. tests results are reported in addition to bleeding (see colored lines in Fig 3b and Methods for the definition of tracking categories and the specification of missing patterns). With time series mimicking the expected tracking behavior of a user whose purpose is to identify their fertile window and pregnancies early on, the accuracy is 93%. The accuracy is of 85% when the tracking behavior is “occasional”, i.e. with an average tracking frequency of about 10% (Fig 3b and Supplementary Material).

The weighted accuracy, *i*.*e*. the accuracy weighted by the uncertainty on the labels at each time-point (see Methods), is higher than the non-weighted accuracy (Fig 3b), which indicates that, as desired, our method provides a higher uncertainty on labels that are different from the ground-truth. Our method is able to warn against potential labeling mistakes.

States recovered with the highest accuracy are the menses and pregnancy states (Fig 3c, Supplementary Materials). The states suffering the most from a low tracking frequency are the states surrounding ovulation. Indeed, without a high tracking frequency, it is impossible to pin-point the day of ovulation. A low accuracy is expected for these states when tracking frequency is low.

### Labeling performance on our real-world dataset

To quantify the performances on our real-world dataset, we used the interactive app embedded in our HiddenSemiMarkov R package to define some ground-truth and label about 11% of our dataset. These labels were independently validated by a fertility awareness methods expert (see Methods and Acknowledgments) and are visualized for the full dataset in the Supplementary Material. Fig 4a provides an example of how a real-world user time series is labeled with our model.

**Figure 4:**
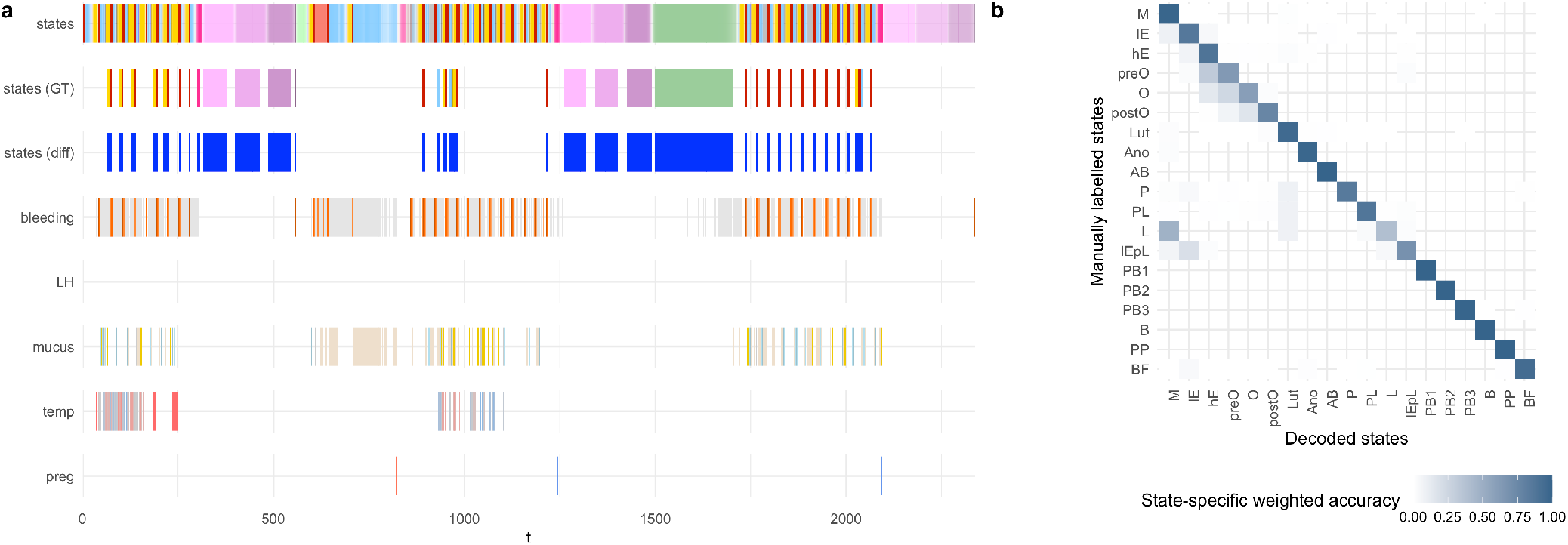
Performances on the Kindara dataset. (a) Example of a time series from a Kindara users with the predicted labels (first row), the manual labels (second row), the difference between the two (third row). (b) Weighted confusion matrix (right) normalized by the number of time-point in each ground-truth state (the sum across rows is equal to 1)

Our results on the partially labeled Kindara dataset are similar to those obtained on simulated data. Menses and pregnancy states are well recovered, while states surrounding ovulation are less accurately recovered (Fig 4b). Here as well, the overall weighted accuracy (94.5%) is higher than the non-weighted one (93%). This also holds true for the state-specific accuracies (Fig 4b, Supplementary Material).

### Predicting the next period

Table 2 provides the mean square error (MSE) on the cycle length prediction for the baseline method (*i*.*e*. average cycle length of the past four cycles) and for our method, using a HSMM fitted to the user’s past four cycles data. The table shows the MSE for predictions done at two different moment of the on-going cycle. The first row provides the MSE when the prediction is made on the first cycle day. The second row provides the MSE when the prediction is made after ovulation, 10 days before the next period. While both method perform similarly at the beginning of the cycle, our method is able to detect ovulation and learn the typical luteal phase duration for that user, providing a much more precise estimate (MSE is 2.88 times smaller) as one progresses through the cycle.

**Table 2:** MSE on the cycle length prediction for both model when made at the start of the on-going cycle or 10 days before the next period.

| Prediction day | MSE Baseline | MSE HSMM |
| --- | --- | --- |
| At cycle start | 17.56 | 16.99 |
| After ovulation (10 days before next cycle) | 17.56 | 6.10 |

## Discussion

Unsupervised labeling of self-reported health records with biologically-relevant states is a challenging problem given the high frequency of missing data, the changes in tracking behavior and the multivariate nature of the data. Here, we presented a hierarchical generative method based on two hidden semi-Markov models. Our results on synthetic data and real-world data, here self-reported fertility body-signs, show accurate recovery of the hidden states sequence. In addition to returning the most likely sequence of hidden states, this framework returns the likelihood at each time-point of this most likely sequence. Our results show that the decoding accuracy is higher when the likelihood is high which implies that our model is able to adequately quantify uncertainty. In contrast, most medical or psychological studies currently use methods which are unable to quantify the uncertainty or the likelihood of their estimates, such as manual labeling or deterministic rules, to identify the timing of reproductive events such as ovulation day. Our method both labels retrospective time series and predicts cycle lengths of ongoing cycles. The method performs 2.88 times better than the baseline method for users with irregular cycles.

Beyond modeling reproductive events, our adaption of hidden semi-Markov models allows (i) for missingness patterns that may differ between variables, *i*.*e*. the probabilities of missing time-points are both state– and variable– dependent and may change over long-term time-scale (ii) for variables of different types (continuous, discrete, categorical) and specified from different marginal distributions and (iii) for partial supervised learning of the model parameters if some ground truth is provided.

We have implemented our method in a publicly available R package (HiddenSemiMarkov) which builds upon the existing implementation of the Viterbi and Forward-Backward algorithms from the mhsmm package (O’Connell & Hojsgaard, 2011).

One limitation of our framework is that within-state dependencies between variables cannot be specified when initializing the model. However, our simulations show that these dependencies are successfully learned when the model is fitted to a sequence of observations which contains these correlations (Supplementary Material). In addition to expected functions of a hidden semi-Markov package, *i*.*e*. functions to specify and fit censored hidden semi-Markov models, simulate time series, and predict sequences of hidden states using the Viterbi or the Forward-Backward algorithm, we also provide several visualization functions for inspecting labeled time series and model parameters. Finally, we implemented an interactive app which can be used to manually label time series and/or confirm predicted labels. This interactive app can be used to create some ground-truth for time series or to use an *interactive boosting* approach to accelerate the fitting process.

This package, in combination with the proposed reproductive model presented here, provide ready-to-use off-the-shelf tools that any scientist interested in studying health and biological variations associated with the menstrual cycle can use. For example, the labeling method presented here can be used to label large retrospective dataset from menstrual cycle tracking app and evaluate the changes in reported symptoms at specific phases of the cycle before and after pregnancies. And, while users in our dataset were naturally cycling, the proposed reproductive model could be extended to allow for the detection of birth-control changes. Therefore, reported symptoms could be compared before and after birth control transitions. Our model will also facilitate the study of associations between the menstrual cycle and the course of chronic conditions (Case & Reid, 2014; Oertelt-Prigione, 2012). Indeed, several studies have already shown that patients which chronic conditions such as inflammatory bowel disease (Bharadwaj et al., 2015), asthma (Ridolo et al., 2019) or systemic lupus erythematosus (Colangelo et al., 2011) report different level of pains or symptoms at different phases of the menstrual cycle. Our framework could thus encourage medical researchers to partner with a tracking app or include a few questions related to participants’ fertility, such as contraceptive use, and their menstrual cycle, such as daily report of their bleeding. This would ensure a more comprehensive understanding of the effect of sex as a biological variable on the course of chronic diseases, the efficacy of treatments or in epidemiological studies.

Beyond the menstrual cycle, the proposed model is ideal for decoding any self-reported time series such as physical activity patterns, or time series of incomplete diagnosis data. As an example from the current pandemic, our hidden semi-Markov model could be fitted to datasets of covid-19 test results and reported symptoms to identify the different phases of infection from “uninfected” to “recovered” over “incubating” and “infectious”. Another application of our hidden semi-Markov model in addition to time series labeling, is the detection of outliers in time series, *i*.*e*. values which may be in the expected variable range of value but that would be unexpected at that particular moment in the time series.

Altogether, this study has demonstrated the accuracy of our coupled hidden semi-Markov models to label multivariate time series with many missing data-points. Our statistical model is especially suited for applications in which the hidden states may impact the frequency of missing data.

## Materials and Methods

### Hidden semi-Markov models

First, we introduce the notation and definitions in the case of a univariate non-censored time series, we then adapt these to multivariate time series, to censored time series and to time series where some ground-truth is available.

### Notation and model parameters

The random variable *X* is measured at a sequence of time-points and may be a discrete, continuous or categorical variable. It is either observed, taking a value *x*, or missing. The hidden state *S* is a discrete, non-observed, random variable whose value *j* is one of the *J* states of the model. We will use the shorthand notation **X** for a sequence of observations of length *N* and define it as an ordered set of *N* values of *X*: **X** = (*x*_1_, *x*_2_, …, *x*_*N*_). A sequence of hidden state is written as **s** = (*s*_1_, *s*_2_, …, *s*_*N*_); *s*_*i*_ or *x*_*i*_ is the shorthand notation for *s*_*t*=*i*_ or *x*_*t*=*i*_.

In general, hidden semi-Markov models are defined by the following set of parameters: *J* is the number of states, *π* are the initial probabilities (*π*_*j*_ = *P* (*S*_1_ = *j*)), *T* are the transition probabilities (*T*_*j,k*_ = *P* (*S*_*t*_ = *j*|*S*_*t*-1_ = *k*)), {*d*_*j*_(*u*)}_*j*=1…*J*_ are the sojourn distributions for each state, *i*.*e*. the distributions of the time spent in a given state (*u* is the relative time variable since the last state transition), and *e*_*x,j*_ are the emission probabilities for each state, i.e. *P* (*X* = *x*|*S* = *j*). This set of parameters is represented by *θ*.

### Likelihood of a sequence of observation and hidden state predictions

The likelihood of a sequence of observations given a sequence of hidden state and the model parameters is given by:

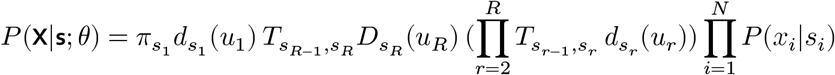

where 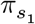 is the initial probability associated with the first state of the sequence, 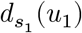 is the sojourn probability of the first state, *u* is the relative time within a state, *r* is an index going through the sequence of state (regardless of their duration) while *i* is an index running along the observation sequence (time-points), *s*_*r*_ is the *r*th state in the state sequence, 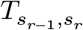 is the transition probability between the state preceding the *r*th state and the *r*th state, 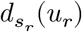 is the sojourn probability of the *r*th state, *R* is the length of the state sequence and 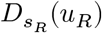 is the “survivor” sojourn probability of the sequence’s last state, *i*.*e*. is it the probability that the state lasts *u*_*R*_ or longer.

Our task is to label time series with a sequence of hidden states. Two algorithms may be used for this purpose: the Viterbi algorithm, which returns the most likely sequence of hidden states, *i*.*e*. the sequence of hidden states that maximizes the likelihood of the sequence of observations, and the Forward-Backward algorithm which returns the probability of each state at each time-point, *i*.*e. P* (*S*_*t*_ = *j*|**X**; *θ*)

Efficient versions of these algorithms for hidden semi-Markov models have been proposed by Guédon et al. (Guédon, 2003) and implemented in C by O’Connel and Hojsgaard (O’Connell & Hojsgaard, 2011). This C implementation is used in our R package, with a minor correction of the backtracking step for the Viterbi algorithm.

### Fitting model parameters to observations

We fit the maximum likelihood estimator and denote it 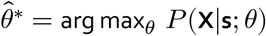

Usually, hidden semi-Markov model parameters are fitted using an *Expectation-Maximization* (EM) approach which iteratively repeats an E-step, in which the hidden sequence of state is estimated and the likelihood is computed given the current parameter values, and an M-step, in which the parameters are updated to maximize the likelihood. These two steps are repeated until the gain in likelihood is smaller than a given threshold.

The Forward-Backward algorithm proposed by Guédon et al. (Guédon, 2003) is used in the E-step to obtain the probability of each state at each time-point. In the M-step, the emission parameters are updated using these probabilities as weights on the observations.

### Multivariate data

In the case of multivariate data, each time-point *i* is associated to a random vector of length 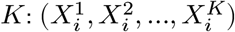. In our case, the first variable is bleeding, the second is mucus, the third is temperature, etc. To adapt for multivariate data, we need to define how the joint emission probabilities at time-point *i, i*.*e*. 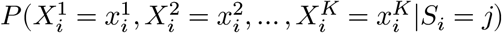 are computed and how potential within-state dependencies between the variables are accounted for.

Past research in reproductive biology has mostly focused on experimentally measuring and describing marginal probabilities. For example, variations in temperature and in cervical mucus have been described separately and there is no available literature to inform us about potential correlations within a particular hormonal state. We thus assume conditional independence of the variables given the hidden state when initializing the joint emission probabilities. Each variable is specified as a non-parametric distribution or by a distribution family and set of parameters. For example, temperature, a continuous variable, may be specified as a normal distribution. Cervical mucus is a categorical variable and may be described by a non-parametric distribution. LH and pregnancy tests results are binary variables (positive or negative results) and may be described as Bernoulli variables.

These initial specifications are used to list all possible observable combination of values and initialize the probability associated with each combination as the product of the marginal probabilities assuming independence conditionally on the states. As the model parameters are fitted to sequences of observations, potential within-state dependencies may be learned as the joint emission probabilities are updated without assuming independence. In the supplementary material, we show how a model is able to learn such dependencies when the direction of the correlation between two variables is the only element differentiating two states. Computationally, within-state dependencies can be learned because continuous variables are discretized into a given number of bins so that all possible combination or variable value can be stored in a table. The number and/or size of the bins can be specified as one of the model parameter.

### Missing data: the censoring model

Self-reported health records are subject to high level of missingness. Typically, the tracking behavior shows high variability between users and within users. The tracking frequency of a user changes over time, both at short term, *i*.*e*. within the menstrual cycle, but also at long term. For example, as users change their reproductive objectives, they may start or stop tracking specific body-signs. Missing observations may be modeled as a two-step process: first, users must open the app on a given day, and second, they must measure and report a specific variable on that day. Both processes can be modeled as a Bernoulli events with state-dependent probabilities which can be specified as parameters: *P* (*X*^1^ = missing, …, *X*^*K*^ = missing|*S* = *j*) = *p*_*j*_ and *P* (*X*^*k*^ = missing|*S* = *j*) = *q*_*j,k*_ for each variable *k*.

Altogether, when a hidden semi-Markov is specified, joint emission probabilities are initialized as:

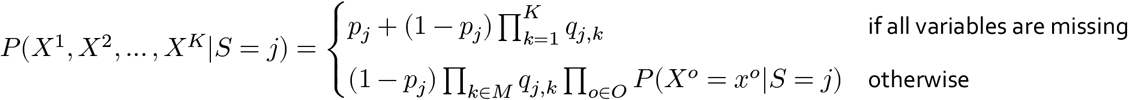

*M* and *O* being the set of missing/observed variables.

These initial probabilities are updated in the M-step of the fitting procedure so that potential dependencies may be learned from sequences of observations.

### Biological interpretations of states: priors on sojourn distributions

Because we want to keep a tight connection between the hidden states and the biological states, we use priors on the sojourn distributions. It would indeed be an undesirable effect of the fitting procedure to get implausible pregnancy duration (*e*.*g*. an 11 months pregnancy). Our framework enables us to specify priors for the sojourns distributions that are used to update the model parameters in the M-step using Bayesian updating:

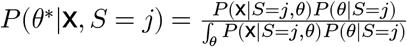 where *P* (*θ*|*S* = *j*) is the prior distribution of the parameters.

The sojourn distributions are updated as follow: 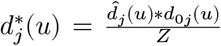 where 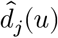 is the sojourn distribution estimated from the Forward-Backward algorithm, 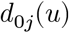 is the prior sojourn distribution specify to match the biological limits of states duration and *Z* is a normalizing factor to ensure that 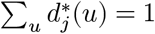

### Semi-supervised learning

Finally, while we do not use semi-supervised learning in the results presented below, we nonetheless wanted our method to be able to leverage information from sparse ground truth. This allows the use of interactive boosting methods which can accelerate the model fitting procedure by confirming decoded state sequences where the model performed well and label with the adequate state parts of sequences where the model did not perform well. Our R package includes an interactive app for manual labeling of decoding validation.

Partial ground truth **s**\* is taken into account in the fitting procedure both in the E-step and in the M-step. In the E-step, the observation probabilities at time-step *i* are adapted as 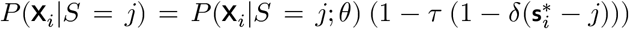
where *δ* is the dirac function and *τ* is a parameter that can be specified and that represents the trust in the provided ground truth. *τ* takes value between 0 (no trust) and 1 (full trust). In the M-step, the posterior state probabilities are adapted as 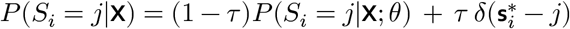.

### Model for reproductive events

To decode our dataset, we defined the simplest hidden semi-Markov model that would as accurately as possible reflect our current knowledge of the menstrual cycle and of reproductive events.

The next paragraphs succinctly describes our model, table 3 provides a description of each state, and figures 2c-d show the model graph and the prior sojourn distributions of most states. The initial marginal emission distributions, as well as a detailed and fully referenced justification for each modeling choice, can be found in the Supplementary Material.

**Table 3:** States of the hidden semi-Markov model of reproductive events

| i | abbr | names | description |
| --- | --- | --- | --- |
| 1 | **M** | Menses | Shedding of the uterine wall, resulting in menstrual bleeding, after an ovulatory or an anovulatory cycle. |
| 2 | **IE** | Early Follicular | Estrogen levels are still low and fertile mucus is not yet observable. |
| 3 | **hE** | Late Follicular | Estrogen levels are rising and fertile mucus is observable. |
| 4 | **preO** | Ovu - 1 | One-day state preceding ovulation to account for a higher probability of a positive LH test compared to previous days. |
| 5 | **O** | Ovu | One-day state during which an egg is released from one of the ovaries. |
| 6 | **postO** | Ovu + 2 | Two-day state during which the mucus becomes less transparent and more sticky and during which the wake-up body temperature increases. |
| 7 | **Lut** | Luteal | Temperature is high (from progesterone), mucus is absent, sticky or creamy and spotting may be reported towards in the last days. |
| 8 | **Ano** | Anovulatory cycle | The anovulatory cycle state follows the follicular phase if a low temperature is consistently observed until the next menses. |
| 9 | **AB** | Anovulatory with bleeding | The anovulatory with bleeding state describes anovulatory cycles in which the users experience quasi-constant bleeding. It follows the menses and the temperature is low throughout this state. |
| 10 | **P** | Implantation (Pregnancy) | The implantation state follows the post-ovulation state. Temperature is high and pregnancy tests may be positive 10-15 days after fertilization (ovulation). |
| 11 | **PL** | Pregnancy with Loss | After implantation, a pregnancy can end in a spontaneous or induced loss. |
| 12 | **L** | Loss | The embryo leaves the uterus, bleeding is usually reported. |
| 13 | **IEpL** | Post-Loss | The post-loss state is similar to the early follicular phase except that pregnancy tests may still be positive (residual HCG in the urine). |
| 14 | **PB1** | Pregnancy with Birth - 1st trimester | Pregnancies ending with a birth are divided into three trimesters. Temperature is very high in the 1st trimester. It has a fixed duration of 84 days (12 weeks). |
| 15 | **PB2** | Pregnancy with Birth - 2nd trimester | Second trimester of the pregnancy, medium temperature. Duration is exactly 12 weeks. |
| 16 | **PB3** | Pregnancy with Birth - 3rd trimester | Thirst trimester of the pregnancy, low temperature. Duration is ~12 weeks, but is variable to account for pre- and post-term births. |
| 17 | **B** | Birth | Birth state is when birth occurs. Bleeding may be reported by users. |
| 18 | **PP** | Post-Partum | After a pregnancy, a mother can either breast-feed or not. If not, the post-partum period is ~ 6-8 weeks before returning to ovulatory cycles. |
| 19 | **BF** | Breast-Feeding | Breast-feeding suppresses the return of ovulation for long period of times. |

We specified a 19-state model composed of 2 main loops (Fig. 2c). The first loop is a 7-state chain describing the successive phases occurring in an ovulatory cycles without conception while the second loop branches out of the first loop after ovulation to describe the successive events following a conception. This conception loop further splits into two sub-loops: one in the event of a pregnancy loss and one in the event of a birth. The birth branch splits into two scenarios depending on whether or not the mother breastfeed their newborn. Breastfeeding typically delays the return of ovulatory cycles. In addition to these main loops, we defined two possible scenarios to capture anovulatory phases The first scenario corresponds to cycles in which quasi-constant bleeding (light or heavy) is reported and in which no signs of ovulation, such as a positive LH test or a rising temperature, would be reported. The second scenario corresponds to cycles in which a low temperature is reported consistently between two bleeding episodes without abnormal bleeding being reported.

All state transitions are uni-directional except for the transition between the “high estrogen” state and the “low estrogen” state. This transition is initialized with a low probability and allows the description of cycles typically experienced by users suffering of poly-cystic ovary syndrome (PCOS). We expect this “hE-lE” transition probabilities to become very low when fitting our initial model on time series reported by healthy users, and to become higher when fitting time series reported by PCOS users or users that experienced delayed ovulation for other reasons.

### Modeling changes in tracking behavior enables decoding of the time series

App users are likely to modify their tracking behavior over time. For example, a user may start using the app simply to track their period, then may completely stop using the app for a few months before starting using the app again, this time with the goal of identifying the days they are most fertile to increase their chances of conception. In each of these three situations, the number of reported variables and their associated missing probabilities are very different. However, the user’s tracking behavior impacts our ability to decode specific reproductive events. Consequently, instead of labeling the whole user’s time series at once, we use a hierarchical approach (see Fig. 2a). First we categorize user’s time series into subsequences with distinct tracking behavior categories, then, if needed, we simplify our reproductive event model to match the information available in each subsequence before predicting their hidden state sequence.

To categorize user’s tracking behavior, we make two assumptions. First, we assume that users are consistent for several months, *i*.*e*. same *p*_*j*_ and *q*_*j,k*_. Second, we assume that they are likely to change their tracking behavior around the time of their period. Periods are ideal boundaries for subsequences as they are the most likely reproductive event that users would report in a menstrual cycle tracking app and they can be considered as true Markovian biological events: the reproductive history before a given period does not affect the future reproductive events. Given these two assumptions, we identify subsequences with specific tracking behavior as follows. We first decode time series of bleeding patterns with a simplified reproductive event model which allow us to roughly estimate timing of periods. Then, we use a second hidden semi-Markov model whose states are associated with specific categories of tracking behavior (Fig 2b). Each category is characterized by a set of reported variables which allows the detection of specific reproductive events. For example, both temperature and pregnancy tests are relevant when identifying pregnancies and differentiating early pregnancy losses from delayed ovulations. The states of this tracking-behavior HSMM are expected to have long sojourns to reflect that a user is likely to keep a similar tracking behavior for a few months. In addition to the tracking-behavior state, we define a *transition* state which lasts only a few days. Any switch between tracking behavior must be done via this transition state whose emission probabilities are specified to coincide with menses as identified earlier (see Fig. 2b). The tracking-behavior HSMM is specified to decode time series that are composed of five binary variables: the first one reflects whether a period was detected by the simplified reproductive model, while the other fours reflects the presence or absence of the four variables (all besides bleeding) that may be reported by the app users.

Once subsequences with consistent tracking behavior have been identified, the reproductive HSMM is slightly modified to match the information contained in each subsequence. These modification consists in one or a combination of three steps. First, unobserved variables are removed from the model emission distributions. Second, if two states (or two loops) may not be distinguishable from the available observations, we set to zero the transition probability for the least common state or loop. For example, anovulatory cycles may not be distinguishable from ovulatory cycles without temperature records; in the absence of temperature, the transition probability to the anovulatory state is set to zero. And third, we initialize to a fixed duration the sojourn distributions of states whose variation in duration can only be infered from the value of given variables. For example, without mucus information, it is irrelevant to attempt to infer the duration of the “high Estrogen” state. Similarly, if a user does not log any variables that would allow the model to estimate the timing of ovulation, variations in the luteal phase duration are irrelevant.

### Datasets

#### Real-world dataset

A de-identified dataset was provided by Prima-Temp (Boulder, Colorado), the company owning the menstrual cycle and fertility tracking app *Kindara*. This study was exempted by the Stanford IRB given the de-identified nature of the dataset. The dataset was lightly pre-processed before being labeled using our hidden semi-Markov models we define in the next section. Temperature reports were transformed into temperature differences from their median value because the interindividual variations in temperature are larger than the within-cycle variations. Specifically, for each user, temperatures marked by the app users as questionable were removed from the time series and their median temperature, computed on the remaining values, was subtracted from their reported values. Additionally, the 13 different possible mucus readings were grouped into 5 categories (none, creamy, fertile, very fertile, sticky) because mucus readings are subjective and the literature is too sparse to allow the distinction between these categories (see Supplementary Material). Finally, because bleeding is the most remarkable body-sign associated with the menstrual cycle, if bleeding was missing on days when other features were reported, it was assumed to have the value “none”.

#### Synthetic dataset

To test our framework and its ability to label real-world time series, we attempted to generate synthetic data that would mimic actual observations. *N*_*u*_ synthetic time series were generated in several steps. First, we adapted our generative hidden semi-Markov model of reproductive events to provide each fictive user with specific menstrual cycle characteristics. Indeed, in the real world, some people have shorter or longer cycles or more variability in their cycle length; other people have noisier temperature measurements. We thus adapted the sojourn distributions or the marginal emission probabilities to reflect these potential differences. Second, we created five levels of tracking assiduity to reflect that some users are tracking more frequently than others. To create these five levels, we first specify censoring probabilities for each state and variables which would mimic the tracking behavior of a diligent user who wish to identify their fertile window and pregnancies early on. We then increase or decrease these probabilities to mimic realistic levels of tracking assiduity (see Supplementary Material). The last step of the data generation process aims at reflecting that (a) users’ tracking frequency may change over time and (b) that not all users track the same set of variable. For example, some users only track their bleeding, while others track their mucus and ovulation tests. We thus divided the users’ time series into an arbitrary number of sub-sequences. Each of these sub-sequences is randomly assigned to a set of tracked features. To simplify the interpretation of the results, 4 groups of realistic sets of tracked features are defined: in the first category, users only report their bleeding patterns; in the second category, their only report bleeding patterns and pregnancy test results; in the third category, they report bleeding, mucus and temperature patterns and in the fourth category, they report all variables. We compute the performance metrics defined above as a function of the tracking frequency and the set of reported variables.

### Labeling performance evaluation

We evaluate the performances of our framework by measuring its ability to recover simulated or manually labeled groundtruth. We compute the accuracy *A* defined as 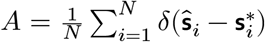 and the weighted accuracy 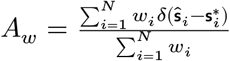 where the weights 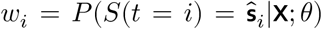 are the posterior state probabilities for the most likely state. We also compute a state-specific weighted accuracy which is defined as 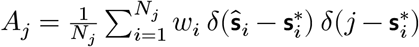

### Predicting the next period date in ovulatory cycles

In addition to labeling user’s time series, our model can also be used to predict the date of the next period. While some individuals have very regular cycles, *i*.*e*. each of their cycle has the same length or little fluctuation around their average cycle length, many individuals have irregular cycles and it may be difficult to predict the length of their next cycle and, consequently, the date of their next period. To evaluate the ability of our method to learn individual-specific characteristics, *i*.*e*. their typical temperature in the follicular or luteal phase or the length of their luteal phase, we selected users in our dataset with highly irregular cycles and compared the predictions from our method with a baseline method which uses the average cycle length of users.

Specifically, we selected stretches of five consecutive ovulatory cycles without pregnancy, fitted our reproductive event model on the first four cycles and made the prediction for the length of the fifth cycles. To predict the cycle length of the fifth cycles, we use the fitted sojourn distributions. To evaluate if the prediction improves as the fifth cycle progresses in time, we perform the prediction from each day since the beginning of the cycle. We decode the fifth cycle up to a given day, detect the last state transition and use the fitted sojourn distributions of the remaining states to predict the timing of the next period from that day. Given that the most variable phase of the cycle is the phase preceding ovulation, we expected our prediction to improve once ovulation is detected. We report the mean square error (MSE) between the predicted and the actual length of the fifth cycle and compare it with the MSE when using the average cycle length of the four previous cycle to predict the length of the fifth cycle.

## Data Availability

Kindara users data are not publicly available. Code for the experiments, supplementary material and synthetic dataset can be found following the Data Availability Link below.

https://github.com/lasy/semiM-Public-Repo

## Acknowledgments

The authors thank the fertility tracking app Kindara as well as their users. They also thank Valentina Salonna, fertility awareness counselor certified by the Symptotherm foundation (Switzerland), for validation of the manual labels on the Kindara data.

## Code and data availability

Code for the experiments, supplementary material and synthetic dataset can be found at https://github.com/lasy/semiM-Public-Repo. Kindara’s users data are not publicly available. The HiddenSemiMarkov R package can be found at https://lasy.github.io/HiddenSemiMarkov/index.html.

